# Perceived heat and rainfall and their longitudinal associations with mental health among the 45+ population in 13 Thai Provinces. A research protocol

**DOI:** 10.1101/2025.08.12.25333382

**Authors:** Romnalin Keanjoom, Sittichai Choosumrong, Keiko Nakamura, Hoang Thuy Linh Nguyen, Nithra Kitreerawuttiwong, Sukrit Kirtsaeng, Juan Gonzalez Hijon, Jacques Wels

## Abstract

**Background:** Climate change has profound implications for mental health, particularly in low- and middle-income countries where environmental exposures and healthcare access vary significantly across regions. Thailand, with its diverse climate zones and rapidly ageing population, provides an important context for exploring the relationship between environmental stressors and psychological well-being.

**Methods:** We will use monthly data from four waves the Health, Aging, and Retirement in Thailand (HART) longitudinal study (2015, 2017, 2020, 2022), covering adults aged 45 and older across 13 provinces, and will combine them with province-linked climate data on monthly temperature, humidity, wind speed and rainfall. To accommodate individual heterogeneity while preserving meaningful regional climate contrasts we will use hybrid mixed-effects multilevel modelling with a within-between decomposition and will estimate the associations between deviations in climate exposure (within individuals over time and average exposure across provinces (between individuals). Climate exposure will include the average monthly heat index (derived from both daily temperatures, wind speed and humidity) as well as the average daily rainfall. The primary outcomes measured psychological distress, mental health and quality of life. Models will successively control for sociodemographic variables, household characteristics, and health status, and include random intercepts at individual level. Multiple imputations will be used to address attrition.

**Expected Results:** The study aims to test whether higher-than-usual perceived heat relative to an individual’s average are associated with higher psychological distress scores, while residing in hotter provinces overall is linked to worse mental health. We will specifically focus on the significance of the interaction between within- and between-person temperature, which would suggest contextual moderation of climate effects.

**Conclusions:** Our findings will highlight the complex interplay between personal trajectories and regional climate experiences in shaping mental health.

## Background

The impacts of climate change extend beyond environmental degradation, significantly influencing mental health through acute and chronic stressors. The direct and indirect pathways through which climate change exacerbates mental health challenges are increasingly recognized ^1^ as many aspects of climate change are susceptible to affect mental health among ageing communities including temperature variability, extreme and abnormal temperatures ^2^. Extreme weather events, such as hurricanes, wildfires, and heatwaves, trigger immediate psychological responses, including anxiety, depression, and post-traumatic stress disorder. Studies have evidenced a relationship between increased temperature and number of suicides and severe psychological distress following extreme weather events ^3^. Several concepts have been used ^4^. The notion of ecological grief is often used to refer to the sorrow experiences because of ecological loss including the loss of species, ecosystems and meaningful landscapes ^5,6^ whilst the notion of eco-anxiety or climate anxiety refers not to the direct impact of climate change but rather concerns about it ^7^.

Social and economic determinants of mental health are also negatively affected by climate change and certain groups are more at risk of climate change and climate change-related hazards, including those with pre-existing health conditions ^1^. Financial inequalities are seen as a mediator of poor mental health when looking at climate change ^3^, deprived communities being more at risk of both poor mental health and environmental hazards ^8^. Deprived communities have lower access to resources, information and protection ^9^ and mental ill health in people exposed to the direct effect of climate change is also affected by policy response. For instance, in the UK, the absence of flood warnings is associated with poorer mental health ^10^. In a meta-analyses, it was pointed out that the impact of climate change on mental health is influenced by gender, socio-economic status, education level and age ^2^.

Furthermore, climate crisis might disrupt health care provision for people with a mental health diagnosis ^3^. On the question of the causal pathways explaining the relationship between climate change and poor mental health, several other factors play a role. The impact of climate change on physical health (e.g., heath exposure of vulnerable people or workers) has potential impacts on mental health ^11^ but effects are also identified at community levels with climate change eroding social environment, thereby potentially affecting mental health, particularly among physically and socio-economically vulnerable people. Sadness, anxiety and concern generated by climate change are also amplified among non-binary and cisgender respondents and a large gap is observed when comparing rural and urban locations ^12^.

Southeast Asia faces compounded vulnerabilities due to frequent extreme weather events and socio-economic disparities. Agricultural and urban populations bear the brunt of these impacts, yet region-specific research on climate-induced mental health challenges remains sparse. Thailand is no exception. Between 1970 and 2017, the average temperature has increased by 1.3 °C causing increasing climate hazards including floods and droughts in many areas of the country ^13^. The severe flooding of summer 2011 resulted in more than 800 deaths, affected 13.6 million people and 65 of the 76 Thai provinces were declared flood disaster zones ^14^. Whilst the magnitude of the flood was spectacular, prolonged droughts decreased agricultural and fishery yields, violent flooding and sea level rise were not new and will likely exacerbate in the coming decades ^15^. Whilst the effect of climate change are strong amongst rural communities and has drastic effects on the agricultural sector ^16^ with framing communities being particularly affected ^17,18^ But cities are not unexposed with frequent flooding in Bangkok ^19^ and increasing cases of seasonal influenza driven by climate change^20^. The geography of Thailand is diverse, with higher precipitations and regular floods in the South and the West of the country and droughts observed in the centre and the east ^21^. Temperatures also very across provinces with lower average temperatures observed in the in the Northwest and particularly high temperatures in the centre of the country (in the North of Bangkok) ^21^.

As climate is diverse across Thai Provinces, it can be expected that its impact on mental health and wellbeing varies by geographical areas. Cross-province comparison of mental health and general health scores is not common in Thailand with most studies focusing on either national scores or some specific provinces. It was estimated that 56 percent of the 60-69 population reported a self-reported health that is fair, bad or very bad ^22^. Age, education, marital status, working status, income, functional status (including activity of daily living – ADL) and chronic diseases contribute to explain poor self-reported health ^22^. Little effect of age and gender on happiness but marital status (being divorced or widowed), how household incomes and having no pay job are associated with strong adverse effects on happiness ^23^. In a study looking at the Songkhla Province, factors associated with poor mental health are age, physical health, presence of a chronic disease, family relationship and participation to social activities ^24^.

Climate change impacts vulnerable populations in specific geographical areas, who face direct exposure and limited access to resources, information, and protection. Both acute and prolonged weather events can lead to immediate traumatic stress or delayed disorders of posttraumatic stress, with effects potentially extending to future generations ^9^.^24^While cross-sectional analyses and ICD-10 data in Thailand have explored associations between climate change, urban heat stress, and mental health ^25,26^. Few studies have longitudinally examined socioeconomic and physical health characteristics as mediators in the relationship between climate variable changes and mental health.

Against this backdrop, this study aims to address three research questions.

[RQ.1] First, to what extent are the differences in mental health explained by climate characteristics attributable to provincial location?

[RQ.2] Second, once a provincial association is established, what portion of the observed change in mental health over the selected period can be attributed to local climate variations?

[RQ.3] Third, are socio-economic and physical health characteristics mediators in the relationship between changes in climate variables and changes in mental health?

## Methods

### Data

**HART** – Health, Ageing and Retirement in Thailand – is biannual household panel survey collected through comprehensive surveys administered to a representative sample of adults aged 45 and over across Thailand. “The Health, Aging, and Retirement in Thailand data is sponsored by Fundamental Fund, Thailand Science Research and Innovation (TSRI) and is conducted by the National Institutes of Development Administration.

The primary goal of the dataset is to gather detailed information on the health status, living conditions, and retirement patterns of the aging population. This includes data on physical and mental health, chronic illnesses, healthcare access, social support, economic resources, and life satisfaction. The surveys also collect information on lifestyle factors such as diet, physical activity, and smoking, as well as data on cognitive function and mental health, including measures of depression and anxiety. The sample for the HART dataset is designed to be representative of the elderly population in Thailand, capturing a wide range of demographic characteristics, including age, gender, education, income level, and geographic location. The sampling method typically involves stratified random sampling to ensure that different regions and socioeconomic groups are adequately represented. Data collection is usually carried out through Computer Assisted Personal Interviewing (CAPI). These interviews are structured to ensure that the data collected is both reliable and comprehensive, allowing for in-depth analysis of the factors affecting aging and retirement in Thailand. HART currently contains four waves, collected in 2015, 2017, 2020 and 2022. The study includes 13 provinces (*Changwat*) from Thailand, covering a diverse range of regions across the country. These provinces span the north, central, and southern areas of Thailand. The dataset features Krabi, located in the south, renowned for its stunning beaches, and Bangkok Metropolis, the capital in the central region. The northern region is represented by Chiang Mai, a cultural and historical hub. Other provinces, such as Khon Kaen, Chanthaburi, Nonthaburi, Pathum Thani, Songkhla, Samut Prakan, Sing Buri, Surin, and Phetchabun, contribute to a balanced geographic representation, showcasing the climate and socio-economic diversity across the north, central, and southern parts of the country.

The data include sensitive information on survey collection points. HART provides monthly information on data collection months allowing to properly address climate values at the time of data collection.

### Mental health and wellbeing

HART includes four variables on self-reported health, quality of life and psychological distress.

<u>Self-reported physical health score</u>: Self-assessed physical health score coded from 1 (poor) to 10 (excellent) in all waves except 2017 where it is coded from 1 to 100. We equivalized the modalities to obtain a scale from 1 to 10 ranging from poor to excellent self-reported mental health. The variable is then rescaled from 1 (excellent) to 10 (poor).

<u>Self-reported mental health score</u>: Self-assessed mental health score coded from 1 (poor) to 10 (excellent) in all waves except 2017 where it is coded from 1 to 100. We equivalized the modalities to obtain a scale from 1 to 10 ranging from poor to excellent self-reported mental health. The variable is then rescaled from 1 (excellent) to 10 (poor).

### <u>Life quality</u>: HART also includes a repeated item on satisfaction with life quality but the answer modalities changed over the waves. It was coded from 0 to 10 in 2015 and 2020, from

0 to 100 in 2017 and from 1 to 10 in 2022. The variable is equivalized from 1 (dissatisfied) to 10 (satisfied). The variable is then rescaled from 1 (satisfied) to 10 (dissatisfied).

<u>Psychological distress</u>: HART also includes several items related to psychological distress, specifically incorporating 10 questions similar to those used in the 12-item General Health Questionnaire (GHQ-12) ^27^, excluding items on feeling worthless and being able to face problems. Items include: feeling bored, lack of concentration, feeling sad, feeling happy, feeling fearful, insomnia or difficulty sleeping, feeling satisfied, feeling lonely, feeling disappointed or unfulfilled and feeling bad, worthless or with no dignity. Response options are based on frequency (1: ’None at all’, 2: ’1-2 days per week’, 3: ’3-4 days per week’, 4: ’5-7 days per week’). After removing NA values, we sum all items (and reverse the happiness variable scale) to calculate a numeric total. The final variable ranges from 1 to 31, with 1 indicating low psychological distress and 31 indicating high psychological distress.

### Meteorological data

We conducted a comprehensive spatial and climatic analysis of Thailand’s weather patterns using meteorological station data and geographic boundary files. The methodology consisted of four key components that were carefully implemented to ensure robust results.

The first component involved station-province matching, where we established precise geographic relationships between weather stations and provincial boundaries. Using Thailand’s administrative shapefiles from the GADM database (version 4.1), we processed spatial joins between station coordinates and provincial boundaries. This required converting raw station location data into spatial points using the sf package in R, followed by point-in-polygon operations to accurately assign each weather station to its corresponding province. Rigorous quality control measures were implemented throughout this process, including the removal of stations with missing or implausible coordinates, and thorough verification of provincial name consistency across all datasets.

For temperature analysis, we processed daily temperature records from each weather station through a comprehensive cleaning pipeline to handle missing values and outliers. Our calculations produced three key metrics: mean daily temperatures derived from averaging available station measurements, identification of extreme temperature events (days exceeding 30 or 35°C), and analysis of annual temperature trends using linear regression models.

Rainfall data processing required special handling of trace precipitation events and missing values. We standardized the dataset by converting all “T” (trace) values to 0.01mm and treating “-” entries as missing data. For each province, we computed multiple rainfall indicators including total annual rainfall through summation of daily precipitation, average daily rainfall amounts, and counts of rainy days (defined as days with >0.1mm precipitation). For the average yearly temperatures, number of hot days (>30) and number of heat days (>35), we selected a minimum threshold of 75 percent data coverage over each selected years with a minimum average stations of 1. For the average yearly rainfall, we select a minimum of 20 percent data coverage and a minimum average stations of 0.50 given the number of days where rainfall information is missing.

Based on these data, we calculate the heat index, or apparent temperature (AT), represents the perceived air temperature by humans, accounting for the combined effects of ambient temperature, relative humidity, and wind speed. While traditional heat indices rely solely on temperature and humidity, Steadman’s extended formula incorporates wind speed to provide a more realistic estimate of thermal comfort in warm conditions^28^. The apparent temperature (AT) in degrees Celsius (°C) is calculated as follow:

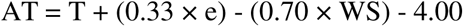

where T is the air temperature (°C), WS is the wind speed (m/s), and e is the water vapor pressure in hPa. The vapor pressure e is derived from relative humidity (RH) (in %) and temperature using the modified Magnus formula:

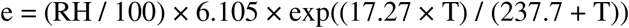

The coefficients 17.27 and 237.7 in the Magnus formula are empirically derived and validated for temperatures between 0–50°C ^29,30^. This formulation reflects how AT increases with humidity (via vapor pressure) and decreases with wind speed, capturing the dual role of moisture and ventilation in perceived heat. This formulation allows apparent temperature to increase with humidity (via vapor pressure) and decrease with higher wind speeds, reflecting the dual influence of moisture and ventilation on perceived heat. Although daily rainfall does not enter the equation directly, it often affects apparent temperature indirectly by increasing near-surface humidity and altering wind patterns following precipitation events. For analytical purposes, rainfall may be included as a contextual variable rather than as an input in the heat index formula itself.

Although daily rainfall does not directly enter the equation, it indirectly influences AT by increasing near-surface humidity and altering wind patterns post-precipitation ^31^. For analytical purposes, rainfall may be included as a contextual variable in heat stress studies rather than as an input to the formula itself.

### Matching HART and meteorological data

HART includes 13 provinces that can be matched with meteorological data. HART includes data collected for the Nonthaburi province but no weather station collected climate information in such a province. We matched the Nonthaburi province with the neighbouring Pathum Thani province that is also included in HART and where a weather station is available. No available climate information was collected for Sing Buri but we used data from the nearby province of Lop Buri. The final dataset includes information from weather stations across 12 provinces throughout Thailand and 13 provinces in the HART dataset. These include Chiang Mail, Khon Kaen, Lop Buri, Phetchabun and Uttaradit in the North; Bangkok, Pathum Thani (including Nonthaburi) and Samut Prakan in the centre; Krabi and Songkhla in the South and Chanthaburi and Surin in the East. Figure 1 shows the HART selected provinces and data collection points and Table 1 shows the study’s 13 selected Provinces, HART 13 provinces and available 12 meteorological stations by province as well as sampling by provincial unit. It can be noted that several provinces include more than one weather stations: 4 stations are available in Bangkok, 3 stations in Phetchabun, Samut Prakan and Surin, 2 in Chanthaburi, Chiang Mai, Khon Kaen, Krabi and Lop Buri and 1 in Pathum Thani and Uttaradit. Province information HART is available throughout the four available waves with a total of 3,708 respondent in 2015, 3,158 respondents in 2017, 3,158 in 2020 and 5,227 in 2022 (including refreshment sample).

**Figure 1.**
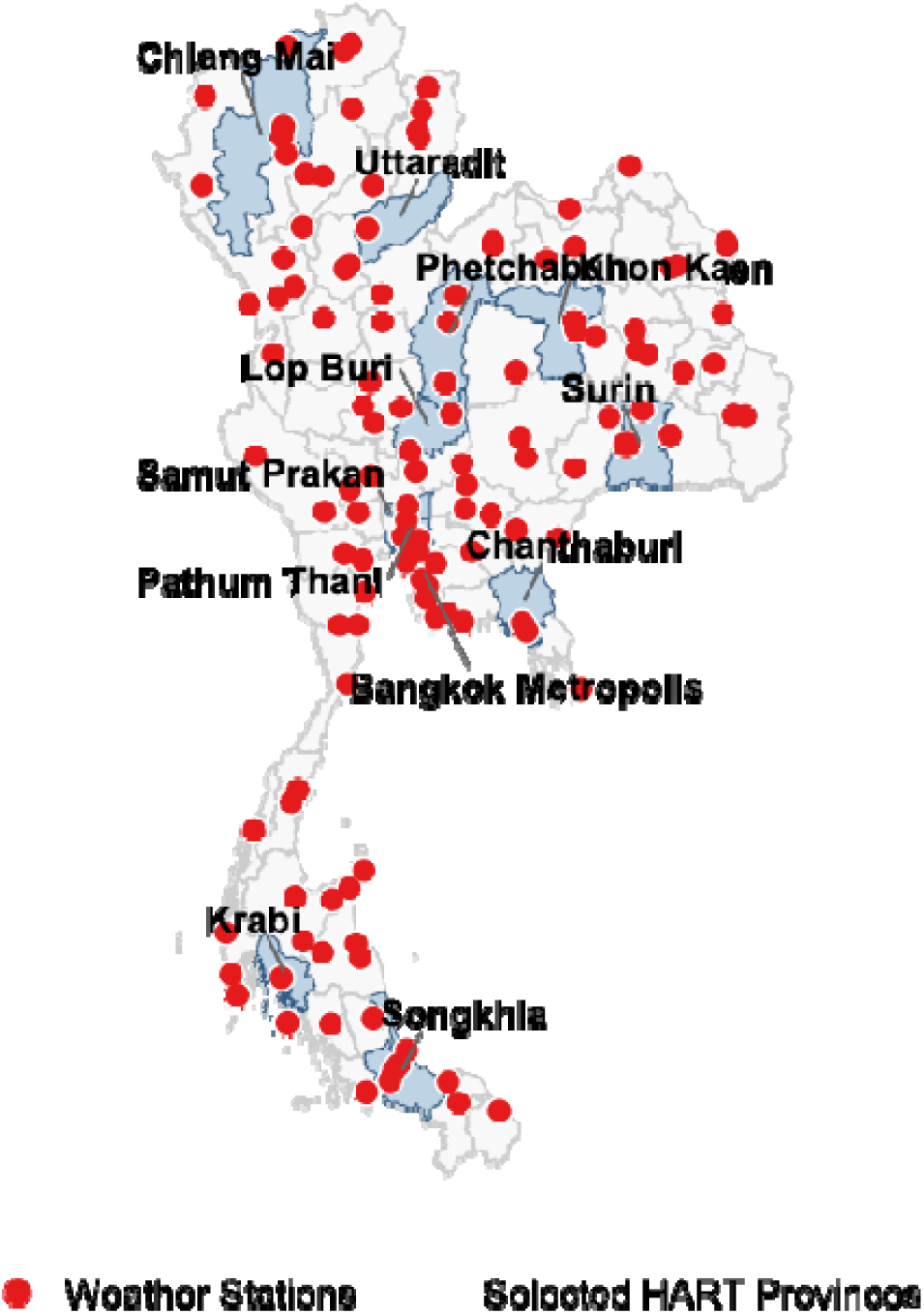
Selected provinces and meteorological data collection points

**Table 1.** Study’s 12 Provinces, HART 13 provinces and available meteorological stations by province.

| Selected Provinces<br>(Changwat) | Health, Ageing and Retirement in<br>Thailand (HART) |  |  |  |  |  |  | Meteorological agency data |  |
| --- | --- | --- | --- | --- | --- | --- | --- | --- | --- |
|  | Available<br>provinces | Localisation | N (valid cases) |  |  |  |  | Available<br>provinces | Weather<br>stations / avg<br>available |
|  |  |  | 2015 | 2017 | 2020 | 2022 | Total |  |  |
| Bangkok | Bangkok | *Centre* | 598 | 288 | 228 | 528 | 1,642 | Bangkok | 5/4 |
| Chanthaburi | Chanthaburi | *East* | 399 | 267 | 209 | 376 | 1,251 | Chanthaburi | 2/2 |
| Chiang Mai | Chiang Mai | *Northwest* | 573 | 474 | 439 | 553 | 2,039 | Chiang Mai | 2/2 |
| Khon Kaen | Khon Kaen | *Northeast* | 623 | 454 | 383 | 549 | 2,009 | Khon Kaen | 2/2 |
| Krabi | Krabi | *Southwest* | 406 | 353 | 278 | 367 | 1,404 | Krabi | 2/2 |
| Lop Buri | Sing Buri | *Centre-North* | 397 | 349 | 291 | 370 | 1,407 | Lop Buri | 2/2 |
| Pathum Thani | Pathum Thani | *Centre* | 198 | 96 | 72 | 168 | 534 | Pathum Thani | 1/1 |
|  | Nonthaburi | *Centre* | 207 | 75 | 42 | 189 | 513 |  |  |
| Phetchabun | Phetchabun | *Centre-North* | 646 | 552 | 509 | 577 | 2,284 | Phetchabun | 3/3 |
| Samut Prakan | Samut Prakan | *Centre* | 198 | 114 | 100 | 192 | 604 | Samut Prakan | 3/3 |
| Songkhla | Songkhla | *South* | 575 | 375 | 339 | 561 | 1,850 | Songkhla | 4/4 |
| Surin | Surin | *East* | 393 | 208 | 197 | 394 | 1,192 | Surin | 3/3 |
| Uttaradit | Uttaradit | *North* | 403 | 103 | 71 | 403 | 980 | Uttaradit | 1/1 |
| **Total** | (13) |  | 5,616 | 3,708 | 3,158 | 5,227 | 17,709 | (12) | 30/29 |

Weather information is available on a daily basis whilst HART data are originally available based on the year of data collection. Access to monthly data information in HART was granted by the Health, Aging, and Retirement in Thailand (HART) project team upon special request.

### Time varying cofounders

The study includes several time-varying cofounders that reflects social determinants of mental health wellbeing found in previous studies ^24,32–35^.

*<u>Activity of daily Living (ADL)</u>*: HART routinely collects information on self-help abilities in dressing, washing the face and/or brushing teeth, bathing and/or washing hair, and eating. Each item is coded across five levels: 1 (’Can perform every step independently without assistance’), 2 (’Needs occasional assistance with some steps’), 3 (’Needs occasional assistance with all steps’), 4 (’Needs assistance every time with some steps’), and 5 (’Needs assistance every time with all steps’). We created a binary variable to distinguish respondents who are fully independent (those who can perform all steps independently, coded as 1) from those who require any assistance (coded as 0).

*<u>Satisfaction with physical health:</u>* respondents to rate their physical health score on a scale from 1 to 100 (in 2015 and 2017) and from 1 to 10 (in 2020 and 2022). We equivalized the scales to get modalities from 1 (poor) to 10 (good)

*<u>Work:</u>* HART collects information on the employment status but complete information on non-work statuses is only available in waves 2017 and 2022. Nevertheless, work/non-work information is collected across waves and we therefore use a binary variable distinguishing respondent in employment or self-employment and those non-employed (ref.).

*<u>Marital status</u>*: repeated information is collected on marital status and is distinguished using two categories: not married nor living together, married or living together (ref.)

*<u>Household composition</u>*: HART includes repeated question on household composition including the following modalities: extended family (ref.), living alone, living with other people (but not relatives), nuclear family (living with a child, with or without a spouse).

*<u>Economic status</u>*: HART collects information on respondents’ satisfaction with their economic status coded on a scale from 1 to 10 (10 indicating poor satisfaction) except in 2017 where the modalities were from 1 to 100. The variable was rescaled for a consistent 1-10 scale estimating the dissatisfaction with current economic status.

### Adjustment layers

We replicated our models using several layers of adjustment. We first provide an *unadjusted model* that only includes climate variables. We then include socio-demographic variables including economic status, household composition, marital status and work status (*socio-demographic adjustment*). Finally, we additionally adjust for ADL and satisfaction with physical health (*fully adjusted model*).

### Modelling

To examine the relationship between climate conditions and mental health, we employed a hybrid mixed-effects regression model ^36^ using the lme4 package in R, also called between-within (BW) model ^37^. This approach allows us to model both within-person (time-varying) and between-region (contextual or time-invariant) effects while accounting for the hierarchical structure of the data, where repeated observations are nested within individuals. By incorporating both fixed and random effects, this model captures both individual-level and group-level variations. Additionally, we included natural cubic splines (from the splines package) to flexibly model temporal trends over the study period. This approach offers greater flexibility than a simple linear trend, allowing for potential nonlinearities in how time affects mental health and wellbeing scores.

The key climate variables, average daily temperature and mean daily rainfall, were decomposed into within-person and between-region components. The within-person component reflects deviations from an individual’s typical climate, while the between-region component represents the regional climate average. This decomposition allows us to examine the impact of short-term, individual-specific climate variations as well as the broader regional climate context. To account for temporal variations, we also incorporated a natural cubic spline for year, with three knots, to adjust for any potential nonlinear temporal trends in the data.

In addition to the climate variables, we included a comprehensive set of time-varying covariates such as employment status, economic satisfaction, marital status, household composition, physical health satisfaction, and limitations in activities of daily living. We also controlled for time-invariant sociodemographic factors, including age, gender, and education level. A random intercept for each individual was included to account for unobserved heterogeneity across participants, capturing individual baseline differences in mental health that do not change over time.

The hybrid mixed-effects model was preferred over traditional fixed effects (FE) or random effects (RE) models because it accounts for both individual-level fluctuations in climate exposure (within-person effects) and the broader regional context (between-region effects) ^38^. One key limitation of fixed-effects models is that they eliminate between-region variation by differencing out all time-invariant factors, including regional baseline climate levels. However, the effect of a climate event, such as a 5-degree increase in temperature, may not be uniform across regions. For instance, an increase of 5 degrees in a region with consistently hot weather may have a different impact on mental health than a similar increase in a cooler region. Fixed-effects models would fail to capture these contextual variations. On the other hand, random-effects models allow for the inclusion of time-invariant covariates such as age and education, but they do not model the within-person dynamics of climate exposure, which is critical for understanding individual-level sensitivity to climate variations. The hybrid approach provides a more comprehensive solution by capturing both individual-specific dynamics and regional contextual effects, thereby allowing for more nuanced and meaningful interpretations of the impact of climate on mental health.

Figure 2 presents a conceptual Directed Acyclic Graph (DAG) illustrating hypothesized pathways linking climate exposure to mental health outcomes within a provincial context. Climate variables – such as temperature, hot days, and rainfall – are theorised to influence wellbeing (mental health scores, psychological distress, and quality of life) through both direct and indirect paths. The indirect pathways operate through two key mediating domains: socio-economic factors (employment, economic status, household composition, and marital status) and health & functioning (self-reported health and activities of daily living).

**Figure 2.**
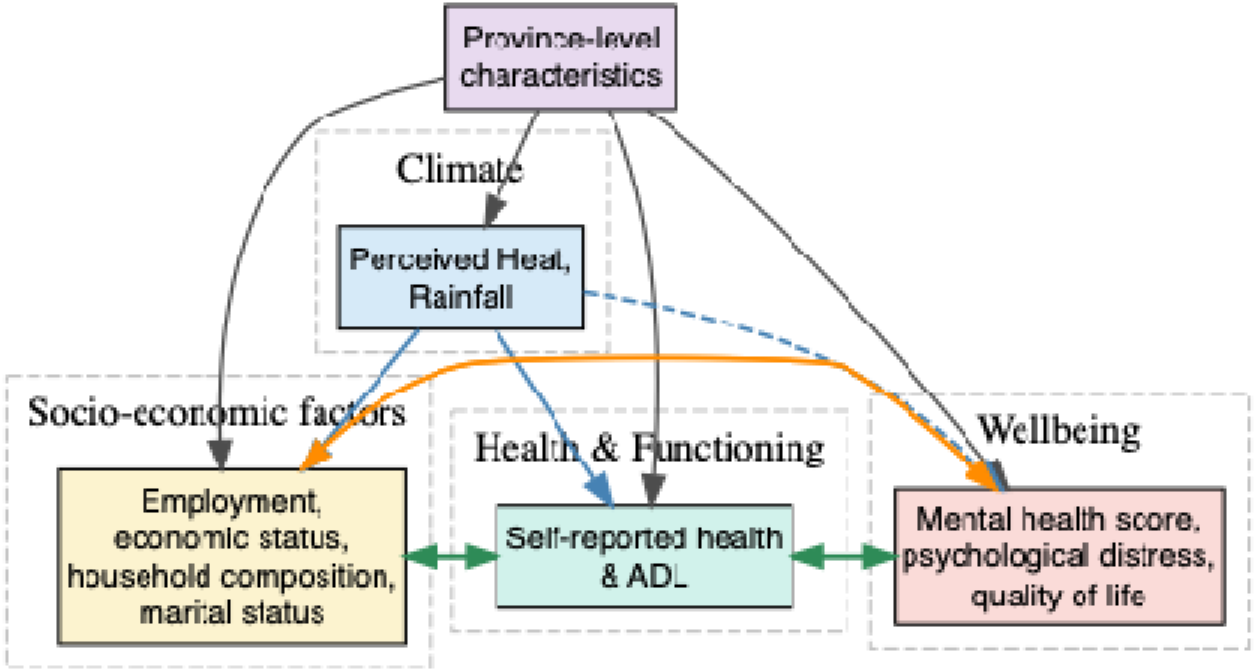
Model specification

Province-level characteristics act as upstream determinants, shaping all clusters and accounting for unobserved time-invariant structural differences across regions. Climate exposure is hypothesized to affect both socio-economic conditions and health & functioning, each of which not only mediates the effect of climate on wellbeing but also interacts reciprocally with one another. These reciprocal relationships reflect the dynamic interplay between social and physical health conditions, wherein, for instance, deteriorating health may constrain economic participation, while economic hardship may erode health and daily functioning. Moreover, both socio-economic conditions and health & functioning are positioned as determinants of mental health outcomes, operating as conduits through which broader environmental and structural factors shape psychological wellbeing. Finally, wellbeing is modelled not only as a dependent outcome but also as a potential influence on socio-economic conditions.

To evaluate potential pathways and address confounding, we implemented a series of models with increasing levels of covariate adjustment. The *unadjusted model* includes only climate variables. The *socio-demographic adjusted model* additionally accounts for time-varying variables related to economic status, household composition, marital status, and employment – capturing potential socio-economic mechanisms through which climate may influence mental health. The *fully adjusted model* further includes self-reported health and limitations in activities of daily living (ADL), which represent the health and functioning domain that may act as both a mediator and a confounder in the climate–mental health relationship. All models cluster standard errors at the provincial level to address within-province correlation and heteroskedasticity, yielding conservative variance estimates.

### Multiple imputations

To address missing data due to attrition across the four selected survey waves, we employed multiple imputation using the Multivariate Imputation by Chained Equations (MICE) package in R. This approach allows for the retention of all available cases and reduces the potential for bias associated with complete-case analysis. We applied Random Forest imputation, a non-parametric method that accommodates complex, nonlinear relationships and interactions between variables without requiring distributional assumptions. The imputation model included all relevant study variables—including outcomes, exposures, and covariates—as well as auxiliary variables predictive of missingness and participation. We generated multiple imputed datasets and pooled results.

### Sensitivity analyses

- Sensitivity check using climate information collected on the previous month (1-month-lag).

## Expected results

The study aims to test whether higher-than-usual temperatures relative to an individual’s average are associated with poorer psychological distress, while residing in hotter provinces overall is linked to worse mental health. We will specifically focus on the significance of the interaction between within- and between-person temperature, which would suggest contextual moderation of climate effects.

## Limitations

This research on climate and mental health is subject to several limitations. First, the characteristics of provinces may evolve over time, but the model does not account for such changes. Additionally, information on household incomes and benefits is not consistently replicated across different survey waves, which may affect comparability. The study also utilizes only three types of climate information, and while it matches data from 13 provinces to 12 climate data regions, this alignment may not be exact, although it is considered representative of Thailand’s climate diversity.

## Data Availability

All data produced are available online at https://hart.nida.ac.th/

## Acknowledgement

This work was partially supported by the Reinventing University Program 2024, The Ministry of Higher Education, Science, Research and Innovation (MHESI), Thailand (Grant number R2567A145). The authors would also like to thank the Thai Meteorological Department (TMD) for providing weather and climate data for Thailand, and the Health, Aging, and Retirement in Thailand (HART) project for providing access to health and retirement data. Jacques Wels acknowledges funding from the FNRS (Belgium) and the UKRI (United Kingdom)

